# Profiling potential targets of anlotinib in human synovial sarcoma patients and immunocompetent mouse models

**DOI:** 10.64898/2025.11.30.25341318

**Authors:** Lara Carroll, Kyllie Smith-Fry, Rahmon Kanmodi, Steve Olsen, Avery Larsen, Brandon Jolley, Linda Morrison, Li Li, Jinxiu Li, Kevin B. Jones

## Abstract

Synovial sarcoma (SS) is an aggressive translocation-driven malignancy characterized by its driving SS18::SSX fusion oncogene. This rare but deadly cancer presents limited treatment options despite an emerging understanding of its dependency on receptor tyrosine kinases (RTKs). Anlotinib is a multi-target tyrosine kinase inhibitor (TKI) that was recently approved in China for treatment of soft tissue sarcomas (STSs), and is now on route through the U.S. clinical pipeline, currently designated an orphan drug. To better understand the therapeutic efficacy of anlotinib for SS, we here investigate the molecular impact of SS on RTK targets of anlotinib. Integrating RNAseq data from STS patient tumors with analyses of our genetically engineered mouse SS models, we performed RNAseq, single-cell transcriptomics, epigenetic profiling, and immunohistochemistry to interrogate a predicted set of kinase anlotinib targets. We identified FGFR1-3 and PDGFRA as the most highly expressed RTKs in SS, targeted both directly and indirectly via enhancer looping by the chromatin-bound SS18::SSX2 oncoprotein. This finding underscores the possibility that rewiring of FGFR (and PDGFRA) signaling—both of which are reported targets of anlotinib—is a fundamental feature of SS. Although *FGFRs1-3* are reportedly targets of anlotinib, TKIs with greater specificity for FGF pathway targets may provide better defense against SS tumor aggression. Our treated SS mice demonstrated only modest attenuation of tumor growth for approximately six weeks, a response that mirrors anlotinib treatment efficacy seen in human patients.

**Significance:** Anlotinib is a small molecule inhibitor shown to inhibit proteins critical to several cancer pathways. We interrogated published human transcriptome datasets and performed preclinical anlotinib trials in an immunocompetent mouse model of synovial sarcoma (SS) to establish a molecular understanding of anlotinib therapy in this rare, but deadly cancer.

## INTRODUCTION

Sarcomas comprise a heterogenous group of malignant neoplasms that arise from mesenchymal tissues, representing approximately 1% of adult cancers, and approximately 15% of pediatric malignancies[1]. Sarcomas arising in soft tissues are generally classified as soft tissue sarcoma (STS), with slightly more than half of all STS patients surviving beyond 5 years[2]. STS is often subcategorized by its genomic complexity, with approximately a third associated with specific, and often singular, genomic derangements generated by a chromosomal translocation.

Synovial sarcoma (SS) is a specific translocation-associated subtype that accounts for 5-10% of all STS. It is the most common non-rhabdomyosarcoma STS in children and the most common STS in the adolescent and young adult population. It is estimated that 1-2 individuals per million population are diagnosed each year in the United States[3]. SS is uniquely characterized by a balanced chromosomal translocation, t(X, 18: p11,q11), creating an oncogene fusing nearly the entire SS18 coded amino acid sequence to the carboxy terminus of SSX1, SSX2, or SSX4[4–6]. This oncoprotein is seen in virtually all cases of SS and has come to define it, with diagnostic confirmation based on identification of the translocation, the fusion oncogene, or the fusion oncoprotein[7]. It is worth noting that SS is not associated with synovial tissues. This misnomer is a remnant from early descriptions of SS among periarticular tumors.

Mutations to receptor tyrosine kinases (RTKs) are highly associated with the development of specific sarcoma subtypes, with elevated activity of RTKs in many other sarcoma subtypes that exhibit tyrosine kinase signaling cascades known to drive proliferation or angiogenesis, making them promising therapeutic targets for this group of cancers[8]. In the 25 years since the introduction of imatinib, the first FDA-approved tyrosine kinase inhibitor (TKI), 8-year overall survival of patients with chronic myeloid leukemia dramatically increased from 6% to 87%[9].

Imatinib was also found to be somewhat effective for treating gastrointestinal stromal tumors and dermatofibrosarcoma protuberans, two sarcoma subtypes driven by mutations that promote hyperactivation of RTKs[10]. In 2012, pazopanib was granted FDA approval for second-line management of STS, with SS proving to be one of the most pazopanib-responsive subtypes in early STS clinical trials[11–13].

Anlotinib (also known as AL3818 or catequentinib) is an orally administered TKI that targets VEGFR1-3, c-Kit, PDGFRA/B, and FGFR1-3[14–18], offering multiple routes to repress angiogenesis (via inhibition of VEGFR2, PDGFRB and FGFR1) as well as direct inhibition of downstream ERK signaling[19]. Anlotinib has additionally been shown to inhibit tumor cell proliferation and promote apoptosis in PDX and cell line models of SS[20]. More recently, anlotinib has been suspected to have some activity against serine/threonine kinases (STKs), as demonstrated by anlotinib’s suppression of Aurora B kinase (AURKB)(14). Currently, anlotinib is approved as a third-line treatment for patients with advanced refractory non-small cell lung cancer and as a second-line treatment for patients with alveolar soft part sarcoma, clear-cell sarcoma, and other types of advanced STSs[21]. Since 2017, 25 clinical trials have been initiated to determine the effects of anlotinib on STSs, with promising results. In a multicenter phase II study, it was demonstrated that several types of STS, including SS, were highly sensitive to anlotinib[22].

Because anlotinib can target a variety of different kinase-dependent pathways, we sought to better understand the molecular impact of SS on potential anlotinib targets. We thus interrogated expression profiles of putative TKI targets exhibiting upregulation in human SS relative to other human STS tumors, and confirmed TKI target expression in our immunocompetent, genetically engineered mouse models of SS (GEMMs), driven by the human *SS18::SSX1* (hSS1) *or SS18::SSX2* (hSS2) oncogene. To address cell specificity of putative TKI target expression within tumors, we analyzed single cell transcriptomics (scRNA-seq) data in untreated SS tumors from hSS2 mice. The therapeutic effect of anlotinib was then tested and confirmed in this mouse model, demonstrating significantly reduced tumor growth for over a month, accompanied by increased tumor infiltration of macrophages in treated adult mice.

## RESULTS

### Human patients and mice show similar SS expression profiles of candidate anlotinib targets

We initially generated a list of 28 potential anlotinib RTK/STK targets in SS, based on published reports from clinical and preclinical trials, as well as *in vitro* studies[13, 23–32] (Table 1). To compare human SS expression levels of each gene with respect to other human STSs, RTK/STK candidates were imported into cBioPortal[33, 34] to generate an expression matrix from TCGA patient data(35) comprising 206 adult STS samples. These samples represent 6 major sarcoma types: 10 SSs, 50 dedifferentiated liposarcomas, 80 leiomyosarcomas, 17 myxofibrosarcomas, 5 malignant peripheral nerve sheath tumors, and 44 undifferentiated pleomorphic sarcomas[35]. **Fig 1A** summarizes STS expression data of each RTK/STK in untreated tumors, with the average kinase expression level of SS (‘SS’) paired with the corresponding expression level averaged across all other sarcomas (‘OS’). Individual kinases are presented on the graph in order of average SS expression levels, with *BRAF* and *FGFR1* representing the lowest and highest expression in SS among the 28 RTK/STKs, respectively. In general, RTK/STK expression levels trend lower in SS than in other sarcomas, including the three canonical VEGF receptors (*FLT1*, *KDR*, and *FLT4*). Notable exceptions are all four fibroblast growth factor receptors (*FGFR1-4*) and *PDGFRA*, which are expressed at relatively higher levels in SS compared to OS in this human dataset.

**Fig 1.**
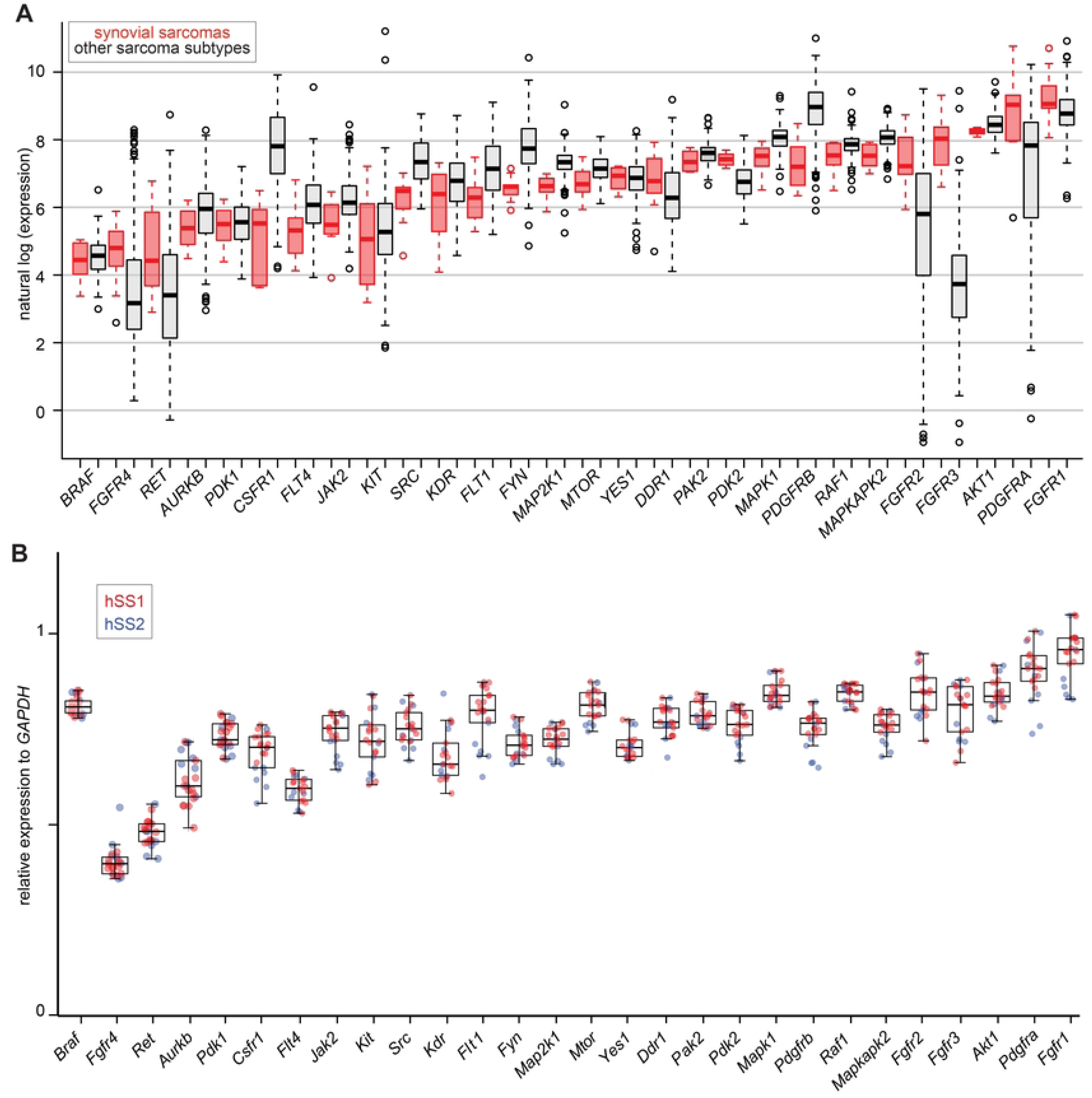
Log transformed expression levels of 28 genes represented as paired box and whisker plots of **(A)** 206 untreated human STS tumor samples (red boxes: synovial sarcoma; white boxes: other sarcomas), and **(B)** untreated mouse tumors driven by human SS18::SSX1 (hSS1, red dots) or SS18::SSX2 (hSS2, blue dots) oncogenes. Gene order reflects average SS expression scores, from lowest SS expression (far left) to highest SS expression (far right). Whisker length encompasses the range of expression values for each gene (excluding outliers in (A), represented by open circles). Box length represents 1^st^ and 3^rd^ quartile for each gene dataset, showing the median score as a solid horizontal line.

We next compared expression levels of these RTK/STKs in untreated tumors from our two genetic mouse models of human SS, driven by induced expression of *SS18::SSX1* (hSS1 model) or *SS18::SSX2* (hSS2 model). RNAseq of whole tumors revealed tight correlations of RTK/STK expression intensities between the two SS tumor types in mice (**Fig. 1B**). Remarkably, SS tumor expression levels in our mouse datasets (presented in the same order as for human SS in **Fig 1A**) generally parallel the human dataset (apart from notable outlier, *Braf*). We thus interrogated our previous hSS2 scRNA-seq dataset(36) to determine cell-specific expression patterns of the 28 RTK/STKs. **Fig 2** shows 21 separate uniform manifold approximation and projection plots (UMAPs), colored by gene expression intensity and superimposed onto the 11 SS projection clusters derived in our original study(36). UMAPs are arranged into four groups based on expression similarity across tumor, endothelial, or macrophage clusters, with two targets (*Pdgfrb* and *Kit*) exhibiting unique expression patterns in fibroblasts and mast cells, respectively. Minimal expression was evidenced for the remaining 7 candidate RTK/STKs (*Fgfr4*, *Ret*, *Pdk1*, *Csf1r*, *Src*, *Mtor*, and *Pdk2*), and representative UMAPs are therefore not presented.

**Fig 2.**
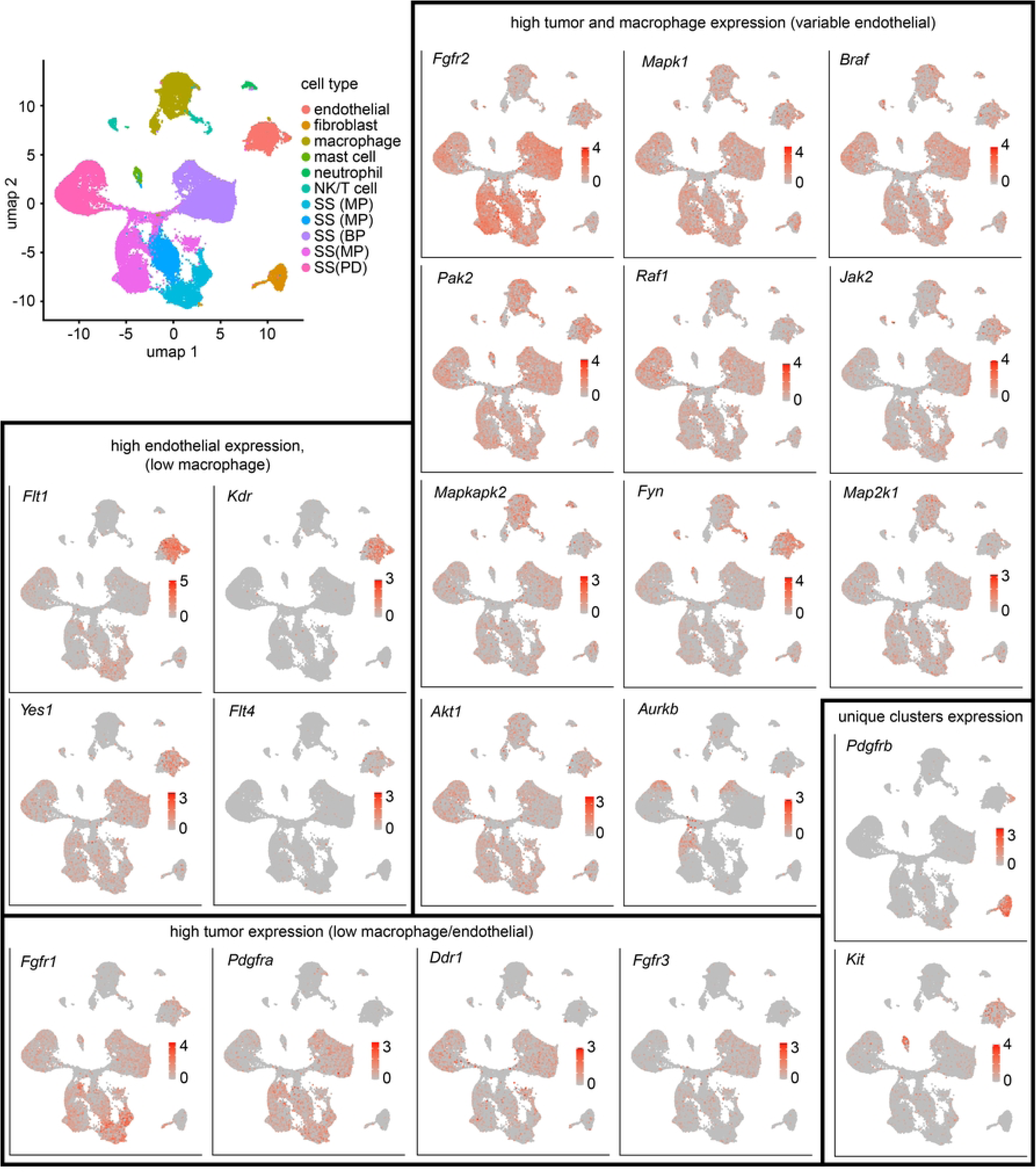
RTK/STK single cell sequencing data of mouse hSS2 tumors presented as UMAPs, with gene expression levels indicated by color intensity and projected onto the 10 clusters defining tumor cell populations (cluster cell types indicated in top left panel; MP = monophasic, BP = biphasic, PD = poorly differentiated). UMAPs are grouped into four general categories based on cluster type-specific expression similarity.

### Epigenetic profiling in GEMMs reveals SS18::SSX2 targeting to promoters and enhancers of highly activated RTKs

Given the prominent expression of *Fgfr1-3* and *Pdgfra* relative to other RTK/STKs in our human and mouse SS datasets, we interrogated ChIP-seq and Hi-ChIP datasets of our untreated hSS2 GEMM tumors[36] to assess the relationship between gene activity and epigenetic regulation of these four genes (along with control genes *Pdgfrb* and *Fgfr4*). Using the Integrative Genomics Viewer (IGV)[37], representative tracks confirm that all four genes are highly expressed, as demonstrated by strong decoration of H3K36me3 marks along the gene body (**Fig. 3A**).

**Fig 3.**
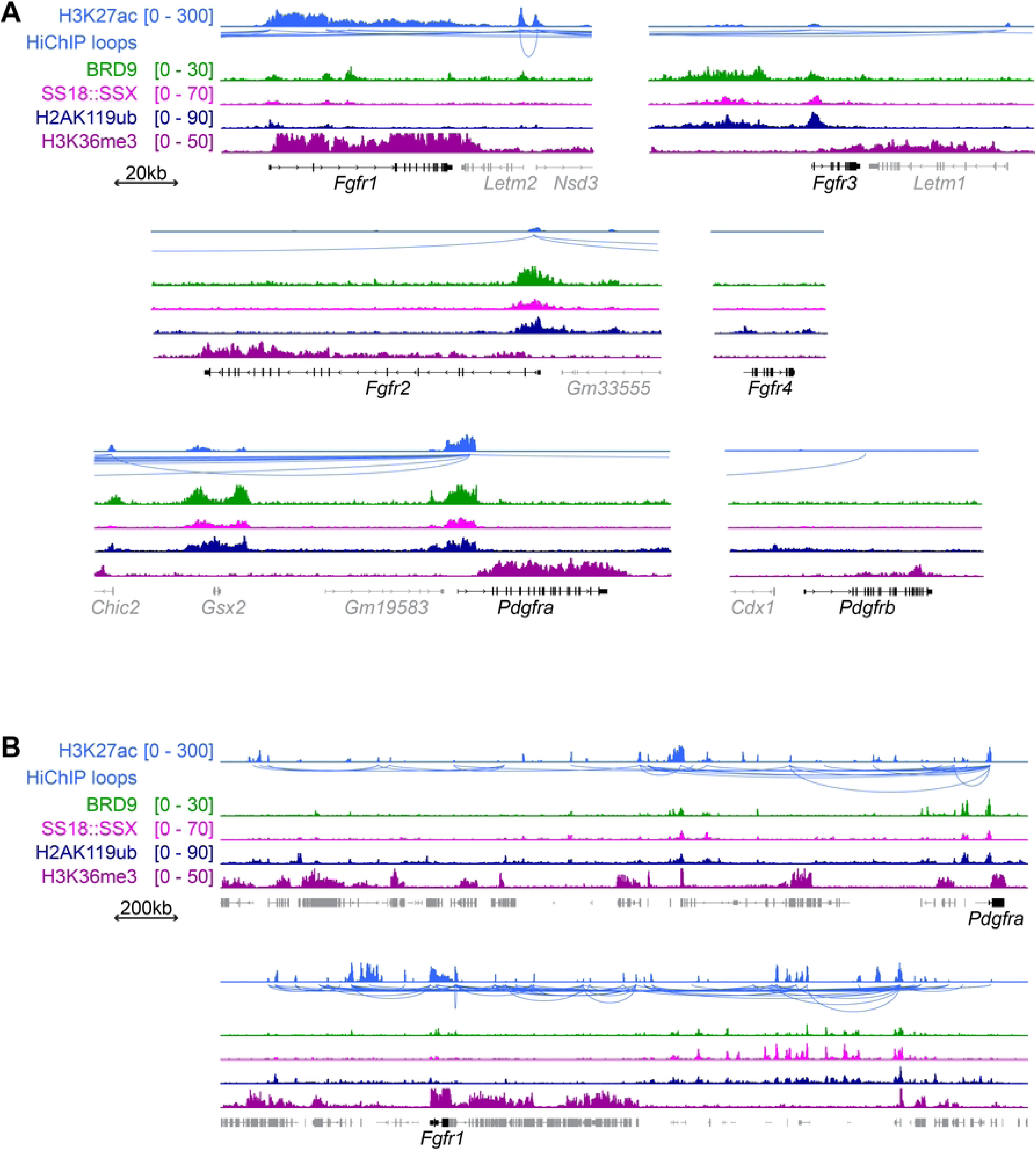
IGV tracks showing epigenetic regulation of *Fgfrs* and *Pdgfrs* in untreated hSS2 mouse tumors (A) *Fgfrs* 1-3 and *Pdgfra*, which were highly expressed in mouse tumors, each exhibiting strong gene-wide decoration by the active H3K36me3 mark, as well as gene occupancy by BRD9, SS18::SSX and H2AK119ub (particularly at promoters), and numerous Hi-ChIP loops from distal enhancers to the promoter. In contrast, *Fgfr4* and *Pdgfrb* gene tracks show minimal active H3K36me3 marks, no SSX::SS18 targeting and a conspicuous lack of distal enhancer looping to either promoter, consistent with low SS expression. (B) Expanded views of *Pdgfra* and *Fgfr1* demonstrate an outsized contribution of superenhancer looping that may amplify gene activation, particularly in the case of *Fgfr1*, which may explain its strong tumor expression despite only modest direct targeting by the fusion oncoprotein and GBAF.

Moreover, multiple distal enhancer loops to each gene’s promoter were evidenced by HiChIP (H3K27ac activity). However, whereas *Pdgfra*, *Fgfr2* and *Fgfr3* promoters are directly and strongly targeted by the SS18::SSX fusion and by the GBAF chromatin remodeling complex (as established by H2AK119ub, SS18::SSX2, and BRD9 occupancy), the *Fgfr1* promoter itself shows more modest SS18::SSX2 and GBAF occupancy. Yet, an expanded view of the *Fgfr1* locus regional chromatin reveals significant looping of very strongly enriched H2AK119ub-, SS18::SSX2- and BRD9-bound enhancers to *Fgfr1* from sites many hundreds of kb downstream, in what appears to be a relative gene desert, suggesting the presence of a fusion-directed superenhancer as the driver of *Fgfr1* transcriptional upregulation in SS (**Fig. 3B**).

*Pdgfra* shows involvement by additional enhancer loops, but not from such strongly clustered enhancers as were evidently looped to *Fgfr1*. As expected from both human and mouse expression datasets (**Fig. 1**), gene tracks of neither *Fgfr4* nor *Pdgfrb* reveal substantial direct or indirect targeting by SS18::SSX2 or by the GBAF chromatin remodeling complex.

### Anlotinib significantly diminishes tumor growth in a mouse model of SS18::SSX2

Unilateral injection of TATCre into the hindlimb of 8-day old mouse pups induces rapid expression of the SS18::SSX2 oncogene in mice homozygous for the hSS2 allele[38] (**Fig. 4A,B**). We previously determined that this protocol allows precise temporal and spatial control of SS18::SSX2 expression, consistently driving growth of palpable tumors within the hindlimb muscle at around 3 months of age. To compare the tumor-suppressive activity of anlotinib in human SS to that in our immunocompetent hSS2 mouse model, we treated mice with established hindlimb tumors (starting at ∼0.5 cm^3^ tumor volume) until endpoint (∼2 cm^3^ tumor volume). Starting soon after treatment onset, anlotinib-treated mice showed a distinct decrease in tumor growth relative to controls (**Fig. 4C**), with a maximal 30% decrease in volume seen 12 weeks after anlotinib treatment initiation, and significant differences in tumor volume maintained between the 8^th^ and 14th week of treatment. By week 14, 50% of the control animals had already reached endpoint, whereas 90% of the anlotinib-treated animals were still alive.

**Fig 4.**
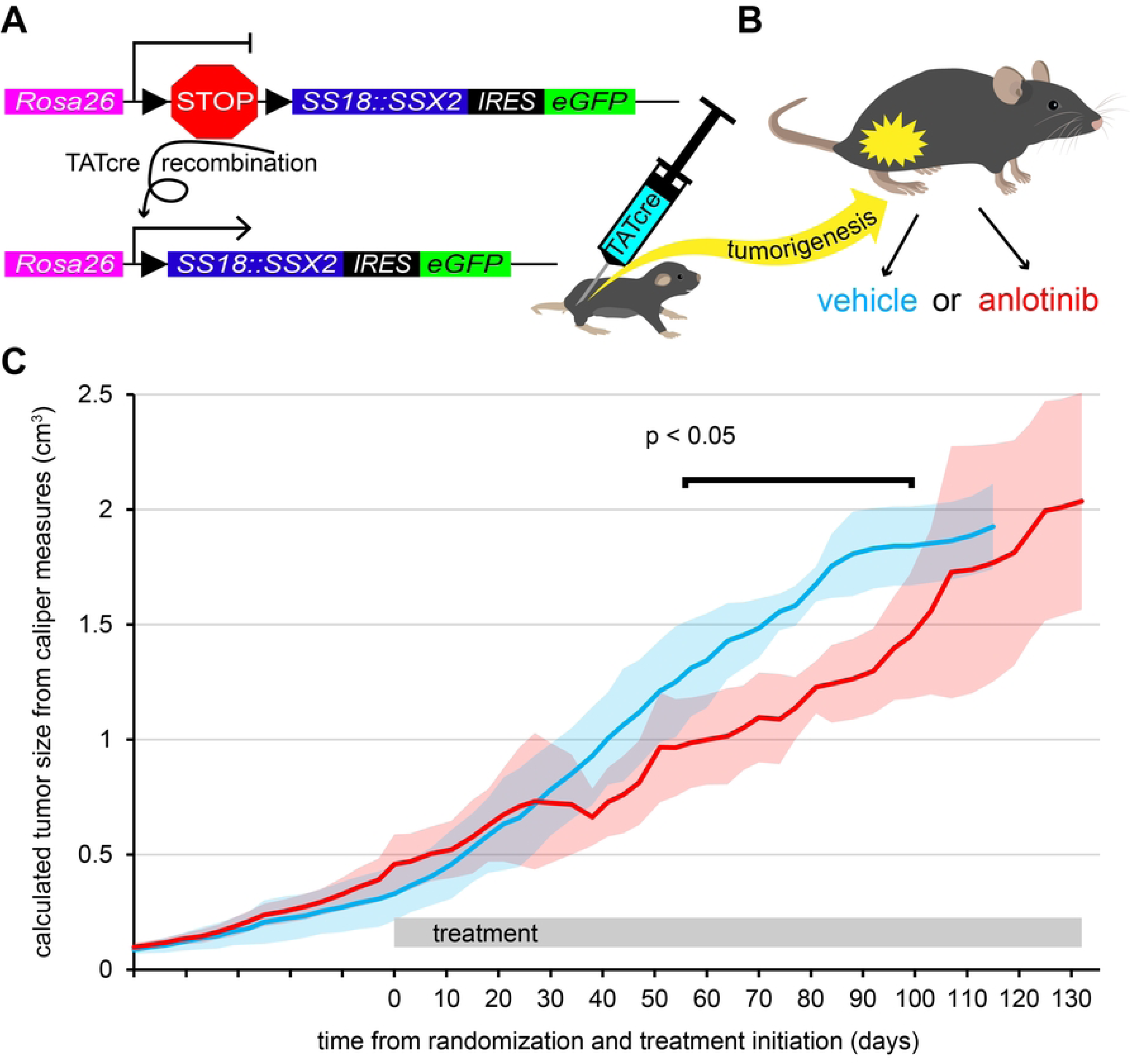
Growth of anlotinib- and vehicle-treated hSS2 tumors in experimental mice. (A) Graphic representation of the conditional *Rosa26-SS18::SSX2-iresGFP* expression construct with loxP flanked stop cassette. Removal of the stop cassette is enabled by hindlimb injection of TATCre at postnatal day 8 (B) with subsequent expression of the fusion gene driving tumor development by approximately 3 months of age. (C) Anlotinib-treated (red) and control (blue) plots of average tumor size ± SD, starting treatment when tumor volume reaches approximately 0.5 cm^3^ and continuing until endpoint when tumor volume reached ∼2 cm^3^.

### Endpoint tumors of hSS2 anlotinib-treated mice show similar SS histopathology and activated ERK and AKT levels as vehicle treated mice, but greater macrophage infiltration

Mouse SS faithfully recapitulates human SS tumors[38–40], with both developing morphologically distinct regions classified histologically as: (a) poorly differentiated, (b) monophasic, or (c) biphasic. Morphologically, these categories differ from one another in cell density, nuclear shape, and the presence/absence of gland-like structures, all of which are histologically distinguishable and may co-occur within a single tumor. Hematoxylin and eosin (H&E) stained vehicle and anlotinib treated tumor sections from our hSS2 mice were evaluated microscopically and scored by a trained expert (blinded to treatment type), with respect to the percentage of each histopathological classification throughout a randomly chosen tumor section (n=9 anlotinib, n=6 vehicle). No significant difference was found between ratios of the three histopathology categories in vehicle- versus anlotinib-treated mouse tumors (**Fig. 5A**), suggesting that histopathological differentiation of mature SS tumors is not influenced by anlotinib treatment. Similarly, no significant histological differences were identified between the two groups with respect to overall cell count or vascular cross-sectional area within sections (**Fig. 5B,C**). In contrast, tumors (n = 5) from anlotinib-treated mice revealed a clear trend of higher F4/80-labelled macrophage infiltration than vehicle-treated (n = 5) mouse tumors (**Fig. 5B,C**).

**Fig 5.**
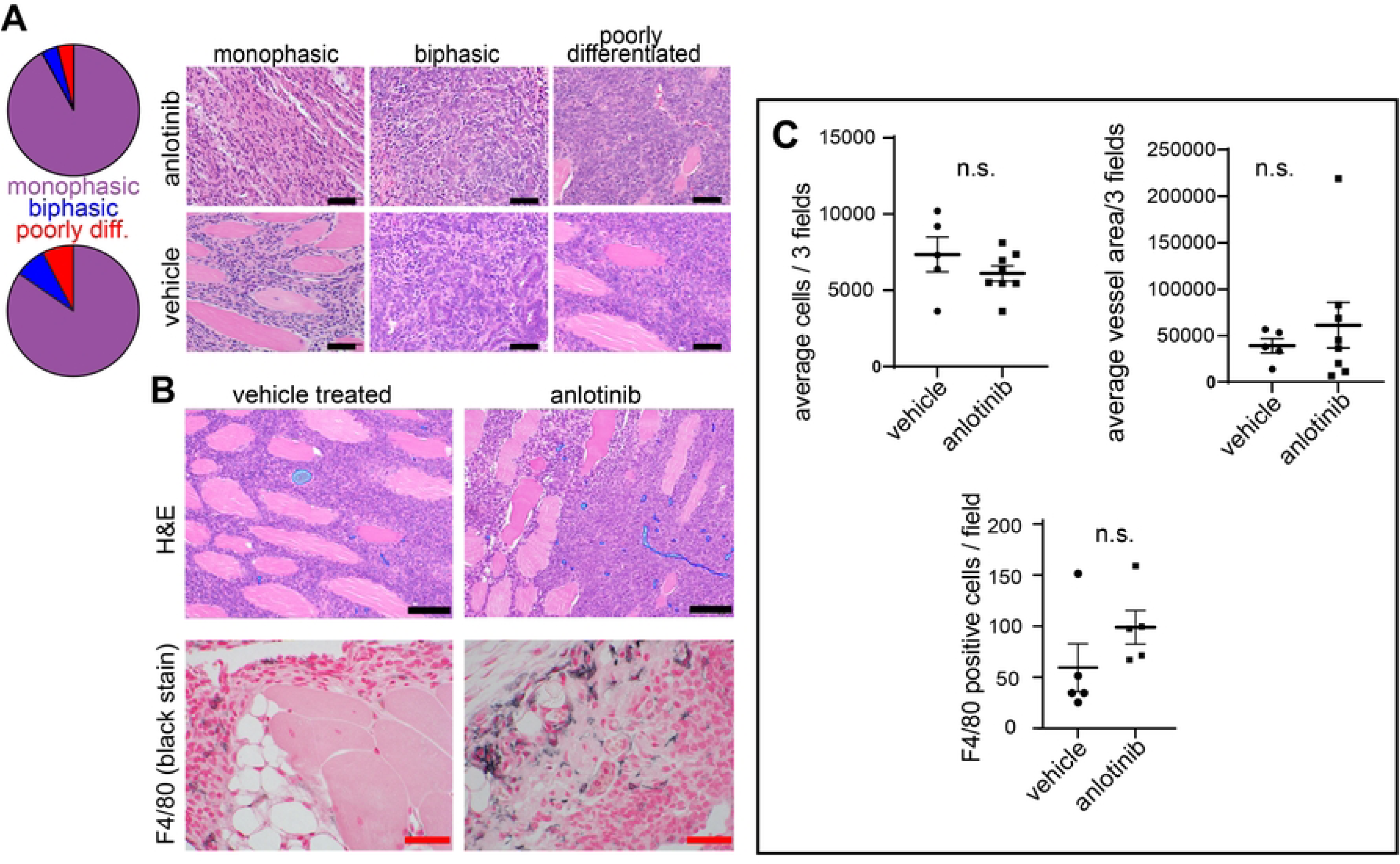
Histopathology and immunology of vehicle- vs. anlotinib-treated hSS2 mouse tumors. (A) Pie charts and representative H&E-stained fields showing proportions of the three typical SS tumor histopathology subtypes (poorly differentiated, monophasic, and biphasic) occurring in mouse hSS2 tumors. Scale bars = 50 µm. (B) Top panels (H&E): representative fields from vehicle- vs. anlotinib-treated tumors showing vessel area (outlined in blue, scale bars = 100 µm). Bottom panels: F4/80 stained macrophages (macrophages stained black with nuclear fast red counterstain, scale bars = 50 µm. (C) Stem and whisker plots showing average ± SEM of vehicle- vs. anlotinib-treated cell counts, vessel areas, and F4/80-labeled cells.

Downstream of VEGFRs, FGFRs and other candidate RTK/STKs, ERK and AKT are critical mediators of cell survival, growth and proliferation. We performed western blots on whole protein lysates of endpoint tumors from hSS2 vehicle and anlotinib-treated mice to quantify phosphorylated (activated) ERK and AKT protein expression (relative to total ERK and AKT protein, respectively) in vehicle- and anlotinib-treated hSS2 mouse tumor samples, reasoning that downstream ERK and AKT activation should be attenuated by anlotinib treatment.

However, considerable variability was found among individual samples both within and between vehicle- and anlotinib-treated groups. Although the average pERK/ERK ratio in tumors from anlotinib-treated mice was 58% lower than that in tumor samples from the vehicle-treated group, this difference was not significant (p = 0.23). Moreover, none of the four proteins tested (normalized to beta-Actin) differed in average expression levels across control and anlotinib groups (**Fig. 6A,B**).

**Fig 6.**
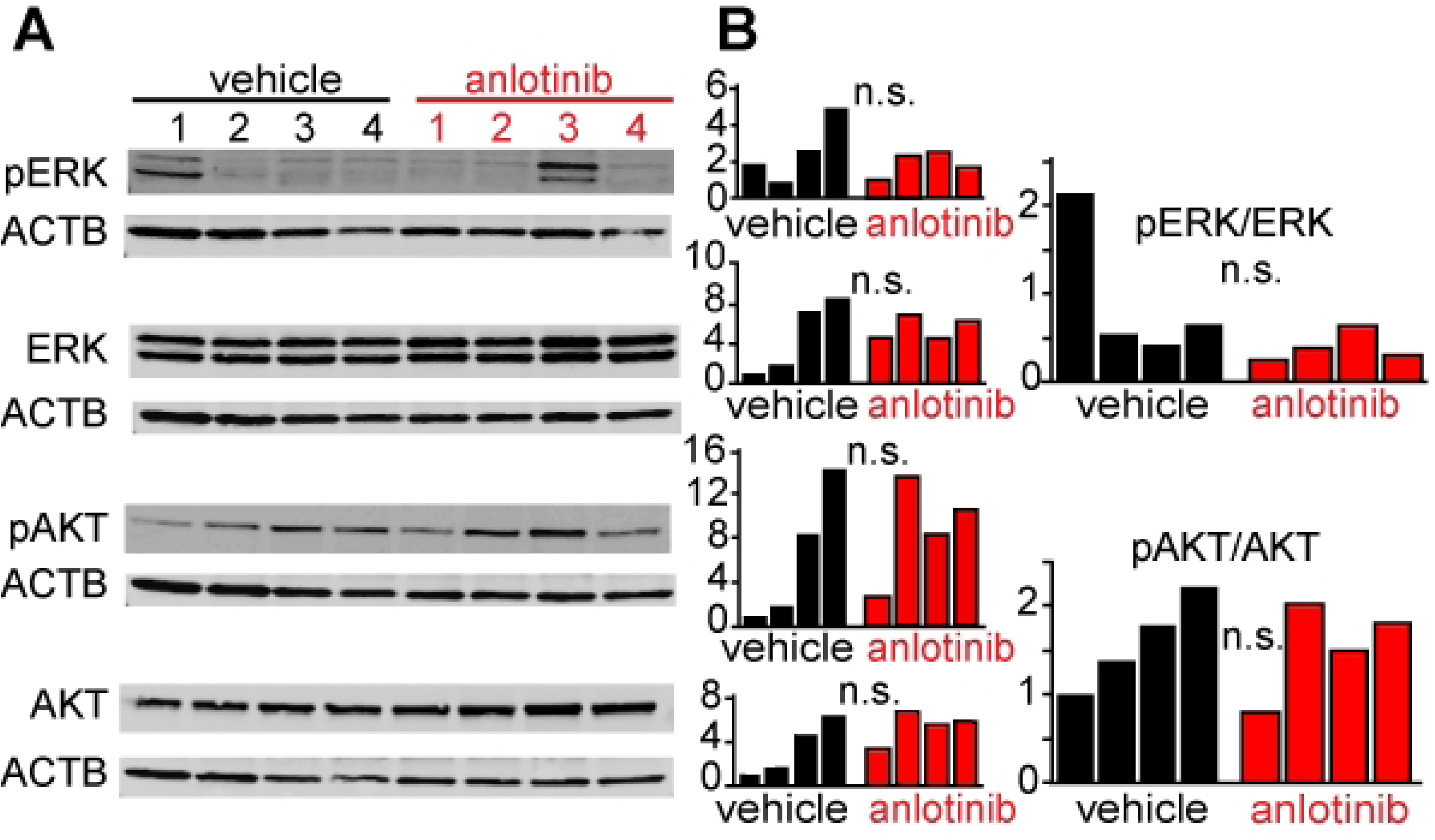
AKT and ERK activation in 4 vehicle- and 4 anlotinib-treated tumors. **(A)** Representative Western blots showing phosphorylated and total ERK and AKT protein bands above corresponding bActin (ACTB) bands. **(B)** Bar graphs show protein quantification of each RTK signal relative to its corresponding bActin signal, normalized to the sample with the lowest expression score (=1). Large bar graphs show phosphorylated/total RTK across all samples.

## DISCUSSION

In further validating our engineered SS mouse models for preclinical anlotinib testing, UMAPs generated from mouse hSS2 scRNA-seq data demonstrate that RTK/STKs are largely expressed within tumor cells, although much expression heterogeneity is present with respect to transcript levels in tumor, immune, and vascular clusters. We identified striking similarities in RTK/STK expression profiles between human and mouse SS tumors. In humans, *FGFR*s 1-3 and *PDGFRa* represent four of the five most highly expressed RTKs, with SS expression levels exceeding the combined averages of ‘other’ STSs (dedifferentiated liposarcoma, leiomyosarcoma, myxofibrosarcoma, malignant peripheral nerve sheath tumors, and undifferentiated pleomorphic sarcoma) for each gene analyzed. *Fgfr1*, *Fgfr2* and *Pdgfra* are similarly the three most highly expressed RTKs in mouse SS, with other RTK/STKs aligning reasonably well to the human dataset. SS reliance on FGFs for sustaining tumorigenesis is supported by independent studies in human cell lines, xenograft and GEMM models[41–43], suggesting that FGF/FGFR signaling is fundamentally rewired in SS. Of the strongly expressed *Fgfrs* in our mouse dataset (as evidenced by extensive H3K36me3 residence along the gene body), *Fgfr1* shows only modest direct targeting by the SS18::SSX oncoprotein, but substantial indirect targeting via numerous oncoprotein- and GBAF-bound enhancer loops, which explains the potent expression of this RTK. In contrast, *Fgfr2*, *Fgfr3* and *Pdgfra* are each driven by a combination of distal enhancer activity and direct promoter targeting by the fusion oncoprotein and by GBAF. Relevantly, we recently showed that SS18::SSX incorporation into GBAF in SS mouse tumors induces GBAF redistribution to promoters and distal enhancers that normally oppose it(36). Combined with the oncoprotein’s direct and enhancer-based targeting of FGFR1-3 chromatin, the markedly elevated expression of these FGFs in SS (with respect both to other RTKs and to other STSs) suggests that direct inhibition of FGFR signaling should be strongly considered for SS therapeutic advancement, particularly as our data suggest that inhibition of canonical VEGF signaling may be less important for SS than for other STSs. Although SS is considered highly angiogenic, expression levels of the three canonical VEGF receptors (*FLT1, KDR, and FLT4*) ranked within the bottom half of surveyed RTK/STKs in the human expression dataset, with each gene trending lower in SS tumors than in other STS subtypes.

Importantly, anlotinib attenuated tumor growth in our hSS2 mouse model (as a ‘first-line’ treatment), to an extent consistent with human clinical trial data. Unlike previous PDX mouse studies, which only examined drug effects following 2-4 weeks of treatment[44–47], we administered anlotinib over a four-month period, demonstrating both long-term tolerability and efficacy. Mice sustained significantly lower tumor volumes for 1.5 months of the four-month treatment duration. However, neither tumor histopathology (tumor classification categories, tumor cell count, vascular area), nor ERK/AKT activity (downstream of putative RTK/STK targets) differed between vehicle- and anlotinib-treated mouse tumors at endpoint. This was largely expected, as continued tumor growth only delayed endpoint volumes by several weeks in anlotinib-treated mice, indicating swift treatment evasion. Nevertheless, the marginally higher macrophage (F4/80 positive) infiltration in anlotinib-treated mice hints at a higher accumulation of dead tumor cells prior to the endpoint harvest (notwithstanding similar tumor cell counts at harvest). It is therefore unsurprising that treated and untreated tumors share similar histopathology. Anlotinib may initially promote apoptosis of tumor cells without measurably affecting tumor vasculature. As anlotinib efficacy eventually ceases, tumor cells likely resume proliferation, leaving endpoint tumors with preserved vasculature and cell counts. Increased macrophage recruitment to sites of necrosis remains as a testament to prior anlotinib activity.

The prospect of a well-tolerated, multi-target treatment for cancer prompted development and approval of several small molecule TKIs over the past two decades, each targeting a discrete subset of VEGFRs, PDGFRs, FGFRs, and other RTK/STKs. These agents, able to selectively inhibit pro-angiogenic and proliferative pathways aberrantly upregulated in cancer, were anticipated to confer less toxicity, better specificity and stronger response rates than traditional chemotherapeutics. Anlotinib, characterized as a potent VEGFR2 inhibitor that additionally targets PDGFRs, FGFRs, cKIT and other RTKs and STKs[17] entered the clinical testing pipeline in China in 2016[31], receiving its first approval for third-line treatment of advanced or metastatic non-small cell lung cancer and second-line treatment of advanced STS[48] shortly thereafter. In August 2025, Anlotinib was approved in China as a first-line treatment for unresectable locally advanced or metastatic STS[49, 50]. This approval trajectory highlights anlotinib’s promise as a therapy for various difficult-to-treat cancers. However, for most STS patients, the benefits of single anlotinib treatment qualify as modest. A recent phase II clinical trial in China[51] found only marginal improvement in progression-free survival (6.8 months) of 40 anlotinib-treated STS patients with advanced disease compared to the reported PFS (4.6 months) following doxorubicin treatment[52, 53]. Unfortunately, the small patient populations of many STS subgroups prevent insufficient enrollment for head-to-head randomized trials comparing anlotinib to other treatments for any specific STS subtype. Moreover, cross-study comparisons are constrained by different dosing schedules, treatment regimes, and outcome measurements. Currently, the reported treatment efficacy of anlotinib appears comparable to that of TKIs preceding anlotinib. However, the superior safety profile of anlotinib merits further exploration for single or combination therapy. Anlotinib is currently designated an orphan drug by the Federal Drug Administration (FDA) for treatment of STS patients in the U.S.A.[50], with a phase III clinical trial (APROMISS, NCT03016819) currently ongoing to evaluate anlotinib in patients with advanced or metastatic alveolar soft part sarcoma, leiomyosarcoma and synovial sarcoma.

## Acknowledgements

We thank Brian Dalley and the High-Throughput Genomics Core at Huntsman Cancer Institute for sequencing support and Tim Parnell and the Bioinformatics Core at Huntsman Cancer Institute for sequencing alignments.

## Author Contributions

K.B.J. conceived the overall experiments. Experiments, data analysis and/or figure preparation were performed by L.C., K.S.F., R.K., S.O., and A.L. The manuscript was written by L.C. with assistance from K.B.J., S.O., A.L., and B.J. All authors reviewed, edited, and approved the final manuscript.

### Ethics statement

All mouse experiments were performed under the auspices of the Institutional Animal Care and Use Committees at the University of Utah in accordance with legal and ethical guidelines.

### Data Availability

Mouse genomics datasets generated in our lab and used in this study are available on the Gene Expression Omnibus: GSE269770 (ChIP-Seq), GSE269772 (RNA-Seq), and GSE269773 (scRNA-Seq).

## METHODS

### Human and mouse RTK/STK expression profiles: RNAseq, scRNA-seq, ChIP-seq and Hi-Chip

Human and mouse sarcoma tumor expression was analyzed for a list of candidate RTK/STK anlotinib target genes curated from published clinical, preclinical and *in vitro* reports(13, 23-32). Human expression levels of these targets were interrogated using data generated by the TCGA research network(35) in cBioPortal(33, 34). RTK/STK expression data of mouse SS tumors (hSS1 and hSS2) were obtained from our previously published RNAseq data(36). Expression data (normalized to corresponding GAPDH expression levels) for both datasets were transformed using the regularized logarithm (rlog) function, ranking genes in order of average human SS expression. For visualization, data frames constructed in R were created for each formatted dataset (**Figs. 1A,B**) to generate a box plot for each gene using the ggplot2 package in R. scRNA-seq data for RTK/STKs was generated using the 10x Genomics Chromium platform (10X Genomics Next GEM Single Cell 3’ Gene Expression Library prep v3.1 with UDI), with raw sequence data processed and aligned to the reference genome mm10 using Cell Ranger software (v7.2.0). Quality control, normalization, and downstream analysis were performed using the Seurat package (v5.0.3) in R as described(36). Dimensionality reduction was carried out using principal component analysis (PCA). Uniform Manifold Approximation and Projection (UMAP) was used for visualization of the clustered cells, implemented through the RunUMAP function in Seurat. The first 20 principal components (PCs) were used as input for UMAP embedding. RTK/STK targets were visualized using the **FeaturePlot** function. ChIP and Hi-ChIP data, visualized by IGV in **Fig. 3**, was originally generated and analyzed as described in our previous publication(36).

### Mouse anlotinib study

Mice used for tumorigenesis studies and anlotinib treatment were generated from parental crosses of our synovial sarcoma mouse line, which carry homozygous conditional alleles of human SS18::SSX2(38-40). SS tumors were initiated by unilateral TATCre injection into the right hindlimb at 8 days of age, driving recombination/deletion of a loxP flanked stop cassette and subsequent expression of the human SS18::SSX2 fusion oncoprotein. Tumor growth was measured with calipers twice weekly starting at approximately 4 weeks of age to accurately time treatment start (tumor volumes estimated as: tumor length x width**^2^**/2), including both males and females from experimental litters in each treatment group. When a tumor reached approximately 0.5 cm^3^, the mouse was randomly assigned to a vehicle control (0.9% saline, 5 mice) or treatment group (1mg/kg anlotinib, Selleck Chemicals, S8726, dissolved in 0.9% saline, 10 mice). Treatments were delivered thereafter 6 days a week via oral gavage, with 2-3 weekly tumor measurements continuing until endpoint, when tumor volume reached approximately 2.0 cm^3^. Human euthanasia was performed when tumor volume reached approximately 2.0 cm^3^, after which tumors were collected for immunohistochemistry, and protein analysis. Despite daily monitoring, 3 animals died prior to sacrifice/necropsy, precluding tumor harvest.

### Histology and immunohistochemistry

Tumor tissues were fixed in 4% paraformaldehyde, dehydrated in increasing ethanol gradients, embedded in paraffin, and sectioned at 4μm thickness for standard H&E staining. For quantitative assessments of histological features in control and anlotinib groups, H&E slides were reviewed (after randomization for order) blind regarding treatment. SS histopathology subtype was classified as previously described(36). Tumor cell number, and vascularity were analyzed in Adobe Photoshop and ImageJ. For each animal, three non-overlapping fields from H&E-stained sections (at 20x obj.) were analyzed. Blood vessels were manually outlined and nuclei were isolated from cytoplasmic using color adjustment in Photoshop. After importing edited images into ImageJ, each was converted to 16-bit black and white, scored for vessel area and total nuclei by a treatment-blind researcher, then averaged for each treatment group.

Macrophage infiltration was analyzed using rat anti-F4/80 antibody (Abcam, 16911). Briefly, paraffin sections were heated to 60°C for 10 minutes, deparaffinized in Histoclear, and rehydrated through ethanol into tap water. Antigen retrieval was performed by boiling sections in citrate-based antigen unmasking solution (Vector Labs, H-3300) for 30 minutes before cooling to room temperature. Following a 30-minute endogenous peroxidase block (0.3% H_2_O_2_ in water) and subsequent PBS washes, sections were incubated for 30 minutes at RT° with 0.5% FBS to block nonspecific proteins. F4/80 antibody, diluted 1:100 in 0.5% FBS, was then applied to each section for overnight incubation at 4°C. The following day, Vector Lab’s Goat anti Rat ABC kit (PK-6104) was used according to manufacturer’s instructions. HRP substrate (Vector Labs, SK-4700) and nuclear fast red counterstain combination were then used to easily visualize black-stained macrophages amidst red nuclei. Macrophage density varied but was generally low across all sections. Therefore, macrophage infiltration (n = 5 mice per treatment) was quantified (blind with respect to treatment) as the average number of macrophages counted across five, 60x obj. non-adjacent regions with the highest microglial density per section and compared by two-tailed Student’s t-test.

### Protein analysis

Protein from snap*-*frozen tumors derived from vehicle-treated and anlotinib-treated hSS2 mice (4 mice/group) was extracted by homogenization in 1x RIPA buffer with 1x protease/phosphatase inhibitor (Sigma, PPC1010). Protein concentration was determined using a Qubit protein kit (ThermoFisher, A50668), with 15ug protein per lane run in precast 10 lane minigels (ThermoFisher) at 100V. Protein bands were then transferred to nitrocellulose membranes at 20V, blocked for 1 hour with Seablock (ThermoFisher, 37527) and incubated overnight at 4°C with primary antibody solution containing both mouse anti-bActin (ACTB 1:10,000, Invitrogen, MA1-140) *and* 1:1000 rabbit anti-pERK (Cell Signaling, 4370T), *or* anti-ERK (Cell Signaling 4695T), *or* anti-pAKT (Cell Signaling, 4060P), *or* anti-AKT (Cell Signaling, 4685S), with antibodies diluted in Seablock. After washing in PBST, membranes were incubated for 1 hour in fluorescent Goat anti-rabbit CW680 (Fisher, NCO252291) and Goat anti-mouse CW800 (Fisher, NC9401841). Following washes, fluorescent signals (bActin and RTKs) were quantified using a Licor Odyssey cLX (LicorBio) imaging system. Signal measurements were imported into excel for analysis, with each RTK signal normalized to its corresponding beta-actin signal. pERK/ERK and pAKT/AKT values for each sample were obtained by dividing each pRTK/bActin signal by its corresponding (total) RTK/bActin signal. Three technical replicates for each antibody were performed per sample.

### Statistics

For the comparison of means between two independent (vehicle vs treatment) groups, we utilized the two-tailed Student’s t-test, as facilitated by GraphPad Prism software (version 9.4.1). We established statistical significance at p-values of less than 0.05.

